# Uncovering clinical rehabilitation technology trends: field observations, mixed methods analysis, and data visualization

**DOI:** 10.1101/2024.03.05.24303809

**Authors:** Courtney Celian, Hannah Redd, Kevin Smaller, Partha Ryali, James L. Patton, David J. Reinkensmeyer, Miriam R. Rafferty

**Author notes:** CORRESPONDING AUTHOR Miriam R. Rafferty, DPT, PhD, 355 E Erie St. (19^th^ floor) Chicago, IL 60611. This study was conducted in accordance with ethical principles and guidelines and was approved by the Northwestern Institutional Review Board (STU00212079).

## Abstract

**Objective:** To analyze real-world rehabilitation technology (RT) use, with a view toward enhancing RT development and adoption.

**Design:** A convergent, mixed-methods study using direct field observations, semi-structured templates, and summative content analysis.

**Setting:** Ten neurorehabilitation units in a single health system.

**Participants:** 3 research clinicians (1OT, 2PTs) observed ∼60 OTs and 70 PTs in inpatient; ∼18 OTs and 30 PTs in outpatient.

**Interventions:** Not applicable

**Main Outcome Measures:** Characteristics of RT, time spent setting up and using RT, and clinician behaviors.

**Results:** 90 distinct devices across 15 different focus areas were inventoried. 329 RT-uses were documented over 44 hours with 42% of inventoried devices used. RT was used more during interventions (72%) than measurement (28%). Intervention devices used frequently were balance/gait (39%), strength/endurance (30%), and transfer/mobility training (16%). Measurement devices were frequently used to measure vitals (83%), followed by grip strength (7%), and upper extremity function (5%). Device characteristics were predominately AC-powered (56%), actuated (57%), monitor-less (53%), multi-use (68%), and required little familiarization (57%). Set-up times were brief (mean ± SD = 3.8±4.21 and 0.8±1.3 for intervention and measurement, respectively); more time was spent with intervention RT (25.6±15) than measurement RT (7.3±11.2). RT nearly always involved verbal instructions (72%) with clinicians providing more feedback on performance (59.7%) than on results (30%). Therapists’ attention was split evenly between direct attention towards the patient during clinician treatment (49.7%) and completing other tasks such as documentation (50%).

**Conclusions:** Even in a tech-friendly hospital, majority of available RT were observed un-used, but identifying these usage patterns is crucial to predict eventual adoption of new designs from earlier stages of RT development. An interactive data visualization page supplement is provided to facilitate this study.

## INTRODUCTION

Rehabilitation technology (RT)^4^ is surprisingly under-used.^5^ Despite the significant funding and years of multi-disciplinary efforts from researchers, developers, and clinicians; cutting-edge tools for individual with disabilities--such as virtual reality,^6^ robotics,^7^ and wearable sensors^8^--rarely see widespread clinical use. The persistent challenge in rehabilitation sciences lies in the disconnect between RT development and clinical use.^5,8–13^ Traditionally, research has focused on efficacy and feasibility studies instead of the factors that influence real-world use of technologies in clinical practice.^13–16^ Perhaps developers do not know enough about the end-stage of this adoption process, leaving a gap to explore the success or failure of effective RT in clinical practice.

To address this gap, we sought to understand the qualities of widely used technologies in clinical treatment, and the factors that facilitate or hinder RT-use. In previous studies, barriers and facilitators of RT into clinical practice have been explored primarily through self-reporting, including surveys,^11,17,18^ interviews,^11,17^ focus groups,^5^ case studies,^14^ and vignettes.^19^ For example, clinicians provided vignettes describing RT-use decisions during treatment sessions, revealing that new RT is not seen as advantageous due to time and complexity.^19^ While these self-reporting methods provide clarity into the implementation challenges and opportunities, they do not fully capture the clinical experience.

Field observations, a form of ethnographic research that uses participant observation to explore practices, provide valuable insights into the practical application of RT in real-world settings.^20^ In the case of RT, it serves as a valuable tool for investigating trends, adoption, and challenges of RT-use in clinical treatment, allowing us to identify commonly used equipment, quantify its use, and elucidate how it is used.

The goal of this manuscript is to enhance RT design and development by understanding observed usage patterns in clinical settings. We did this by observing clinical RT-use and inventorying RT available in inpatient and outpatient settings. We sought to describe (1) the characteristics of RT associated with therapist use; (2) how much time therapists spend using RT; and (3) therapists’ behaviors surrounding RT-use. We intended to build on our previous vignette study, aiming to understand clinicians’ on-the-spot decision-making regarding the use or non-use of RT.^19^

## METHODS

### Study Design

This study was conducted in accordance with ethical principles and guidelines, approved by the Northwestern Institutional Review Board (STU00212079) with a waiver of informed consent from observed patients or clinicians. Given the observational design of the study, no masking or randomization was used. Clinicians followed routine clinical practice with no added treatments or assessments. No patient information, protected health information, or identifiers were documented. The director of the allied health facility authorized all observations. A flyer containing information about the study was sent to the sites’ managers via email to inform their team of the researchers’ presence during observation days.

### Context and sample

The field observations and inventory were conducted in our rehabilitation health system, a flagship translational research hospital designated as a national rehabilitation innovation center.^21^ Thus, there is ample RT in all clinical spaces increasing therapist access and opportunity for use. The health system includes adult inpatient treatment on 7 floors within the flagship hospital, 5 outpatient interdisciplinary DayRehab^©^ sites, and 3 outpatient neurologic rehabilitation clinics. Data collection occurred in 10 patient units across 3 different settings: adult inpatient treatment floors (7 inpatient units), one DayRehab^©^ site (2 units) located adjacent to the flagship hospital, and one outpatient site (1 unit) located within the flagship hospital building. The study observed approximately 70 Physical Therapists (PTs), 60 Occupational Therapists (OTs), and 35 Speech Language Pathologists (SLPs) employed at the inpatient rehabilitation facility; 13 PTs, 12 OTs, and 10 SLPs at the DayRehab^©^ outpatient location; 17 PTs, 6 OTs, and 4 SLPs employed in outpatient neurologic rehabilitation clinic. Even though clinical treatment occurs in a variety of different areas around each setting (e.g., inpatient rooms, hospital hallways, private treatment rooms, and shared gym spaces) we contained the observations and inventory to only shared gym spaces.

### Data Collection

Two trained clinician research team members (CC, HR) made direct field observations^2^ using a semi-structured template in Excel ^a^, and inventoried each clinical treatment gym in all 10 patient units. This included a range of RT for gait (e.g., overhead gait tracks and treadmills), assessment tools (e.g., upper extremity assessment kits, dynamometers), and general rehab equipment (e.g., adjustable mat table and parallel bars). We excluded simple items such as towels, canes, and walkers as they are used as tools in rehabilitation but not specifically designed as RT for therapists. Data collected in the field observations included descriptive (e.g., RT observed used, instructions given to patient), categorical (e.g., clinician type), and quantitative (e.g., time spent setting up RT) variables. Only descriptive variables (e.g., RT) were recorded in the inventory. Table 1 contains operational definitions and examples of all qualitative and quantitative data.

**Table 1:**
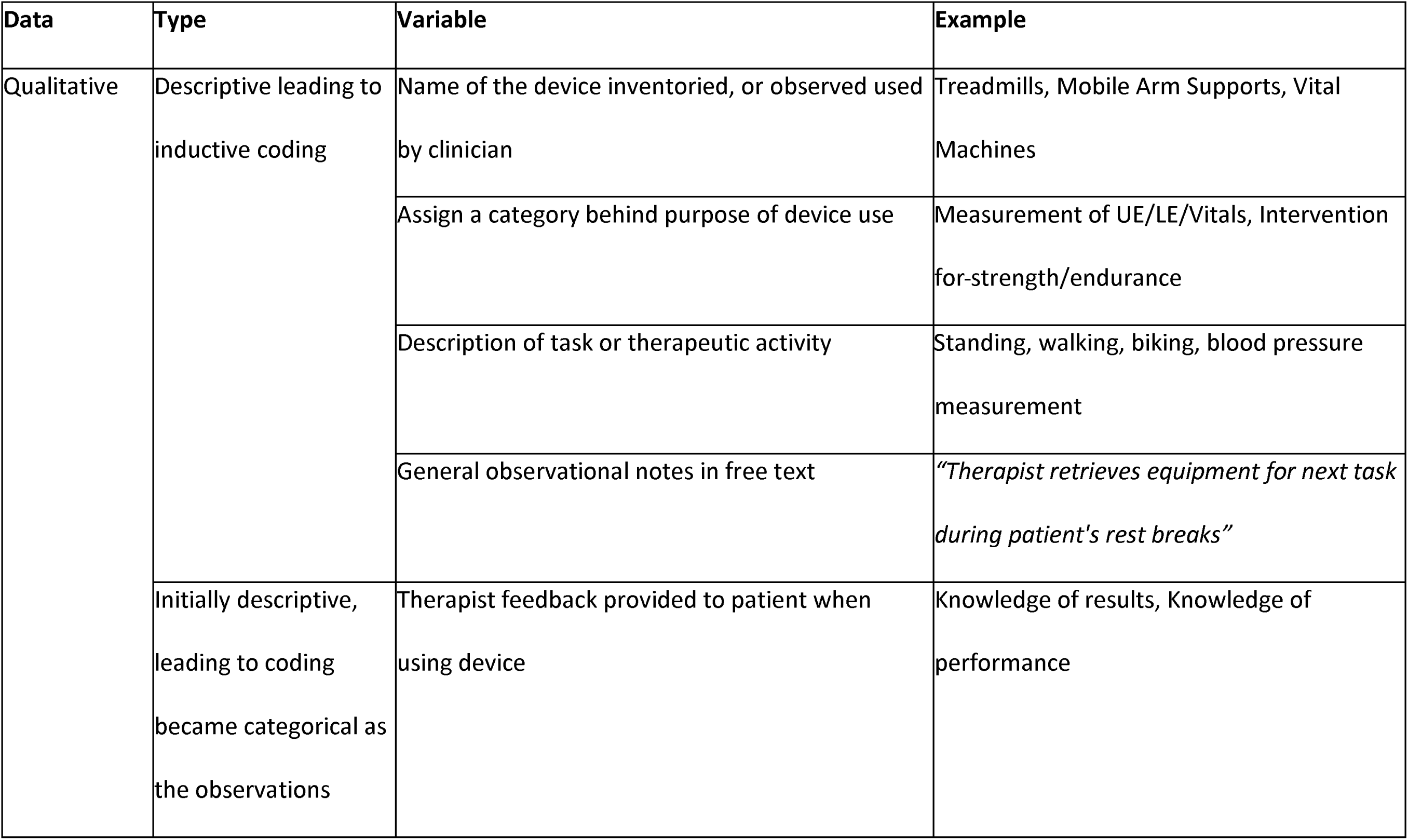
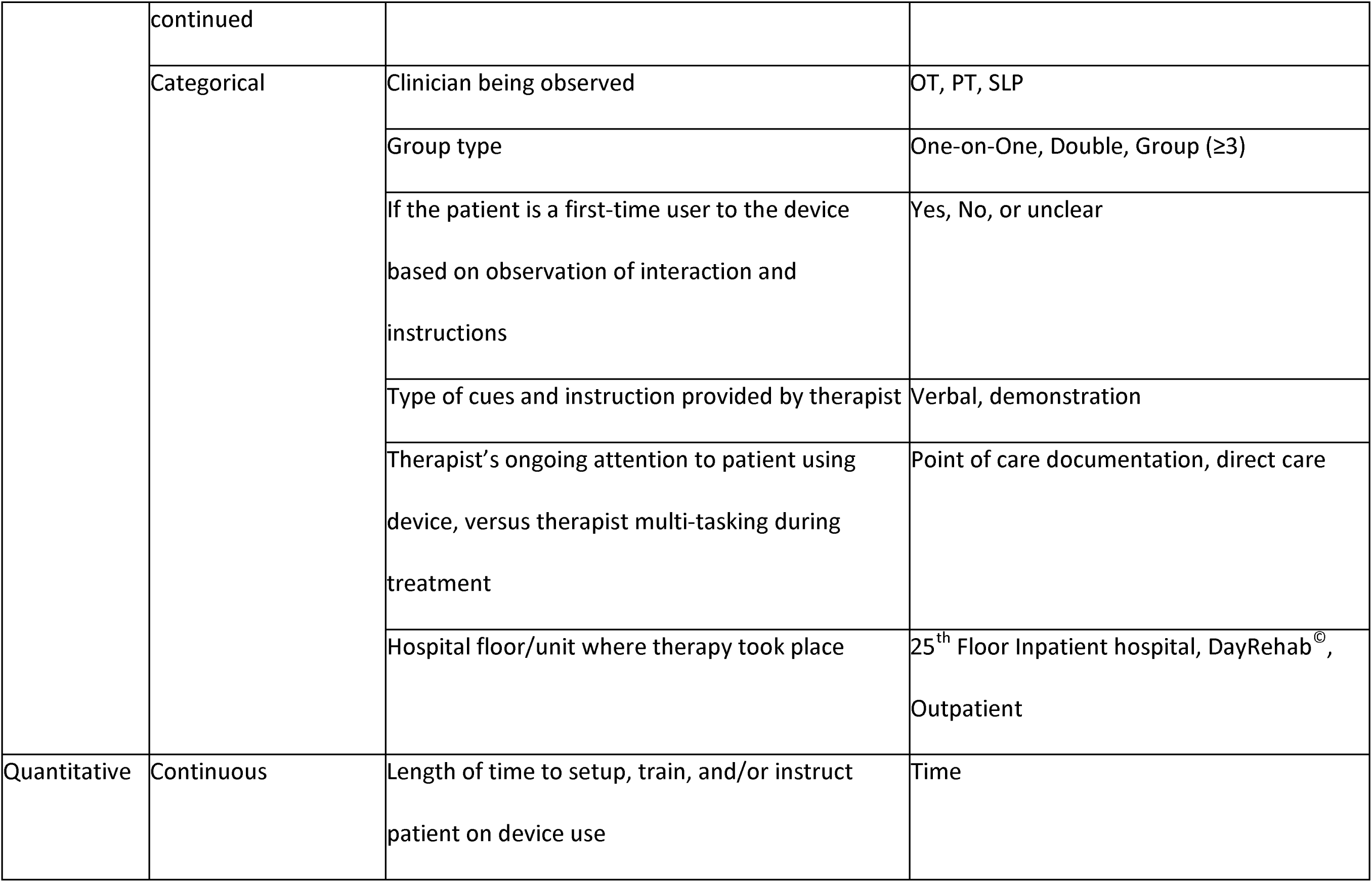
Qualitative and quantitative data collected in field observations and inventory. Table includes the type of data collected, distinguishes each variable, and provides an example.

### Analysis

A convergent, mixed methods design^1^ used conventional reporting guidelines.^22^ Two coders (CC,HR) refined descriptive variables through inductive coding, communicated regularly to resolve discrepancies, and a third coder (MR) assisted in consensus building on the coding strategy. We used qualitative content analysis, using a summative approach,^3^ to analyze the descriptive variables; this resulted in the creation of multiple subcategories under the different types of RT. We used descriptive statistics to analyze categorical data (e.g., clinician type, feedback type, etc.), representing the results through counts and percents. Data integration occurred through transforming qualitative data into quantitative, using content analysis.^23^ We analyzed time quantitatively as a continuous variable measured in minutes. We analyzed and visualized data using Excel^a^ and R software^b^, and used Python^c^ for exploratory data analysis to investigate the most prevalent combination of RT characteristics used during clinical treatment, and understand the plurality of RT.

## RESULTS

Three trained research clinicians (1 OT, and 2 PTs) completed field observations intermittently between January 2021 through September 2022.They observed for over 44 hours in 60 to 190-minute increments throughout the day until reaching data saturation (defined as the lack of new RT, or RT used in novel ways). The clinicians completed over 24 hours of observations in the inpatient setting, over 10 hours in DayRehab^©^, and over 9 hours completed in outpatient. In total, they observed 329 RT-uses. They did not observe SLPs using RT in the shared gym space; therefore, the analysis focuses on OTs and PTs.

Two trained research clinicians (1 OT, 1 PT) inventoried 286 RT across all settings; after resolving duplicates, the total inventory revealed 90 distinct devices across 15 different focus areas, with 42% (n=38) observed used (supplemental Table 1). Of all inventoried devices, 21% (n=19) were categorized as gait and/or balance focused, with 36% (n=7) observed used; 13% were for strengthening and/or endurance (n=12), with 75% (n=9) observed used. There are 6 device focus categories in which there are only 1 or 2 device models in the inventory (e.g., vitals, cognition). See supplementary information for a link to an interactive Sankey Diagram that explores the relationships between the inventoried devices.

### Field Observations

Inductive coding of the field observations revealed two RT-use activities: intervention and measurement. Intervention involves actions by clinicians to create, promote, restore, maintain, or modify function, while measurement assesses initial performance or outcomes.^24^ Table 2 details categories and subcategories of observed RT, with counts and percentages. Supplementary materials include an interactive pie chart and Sankey diagram illustrating the proportion of RT characteristics in intervention and measurement RT from the field observations (supplemental Sankey diagram 1).

**Table 2:** Characteristics of clinicians, treatments, and rehabilitation technologies (RT) observed used in field observations. Table includes definitions, counts, and percentages by category and sub-category, separated by type and total.

| **Category** | **Sub-Category** | **Definition** | **Intervention**<br>**RT**<br>**(237, 72%)** | **Measurement**<br>**RT**<br>**(92, 28%)** | **Total**<br>**(329)** |
| --- | --- | --- | --- | --- | --- |
| **Clinician**<br>**Type** | OT | Occupational Therapists | 56 (24%) | 14 (15%) | 70 (21%) |
|  | PT | Physical Therapists | 173 (73%) | 73 (79%) | 246 (75%) |
|  | Not Documented | -- | 8 (3%) | 5 (5%) | 13 (4%) |
| **Treatment**<br>**Type** | One On One | Clinician working with 1 patient | 169 (71%) | 78 (85%) | 247 (76%) |
|  | Double | Clinician working with 2 patients simultaneously | 20 (8%) | 1 (1%) | 21 (6%) |
|  | Group | Clinician working with 3 or more patients simultaneously | 13 (5%) | 1 (1%) | 14 (4%) |
|  | Not Documented | -- | 35 (15%) | 12 (13%) | 47 (14%) |
| **Power**<br>**Source** | Electrical Power | Powered by alternating current (e.g., wall plug/electricity) | 183 (77%) | 1 (1%) | 184 (56%) |
|  | Battery Powered | RT powered by a battery source, also includes rechargeable batteries; or does not remain plugged into a wall outlet to be powered | 19 (8%) | 81 (88%) | 100 (30%) |
|  | Unpowered | No source of power; or uses cables, pullies, or pneumatics to operate | 35 (15%) | 10 (11%) | 45 (14%) |
| **Smart RT** | Actuated | Powered RT, motor, moves | 185 (78%) | 1 (1%) | 186 (57%) |
|  | Unactuated | Typically unpowered, does not move | 52 (22%) | 91 (99%) | 143 (43%) |
| **Display** | Monitor | Visual display screen | 75 (32%) | 81 (88%) | 156 (47%) |
|  | No Monitor | No visual display screen | 162 (68%) | 11 (12%) | 173 (53%) |
| **Versatility** | Multi-Use Tool | RT that can be used for multiple types of interventions, and/or measurements. | 222 (94%) | 2 (2%) | 224 (68%) |
|  | Single-Use Tool | RT that serves only one function | 15 (6%) | 90 (98%) | 105 (32%) |
| **Training Level** | Entry-Level | Taught in clinical schools, no additional training beyond OT/PT degree | 109 (46%) | 78 (85%) | 187 (57%) |
|  | Short | 1-2 hours of additional training | 111 (47%) | 10 (11%) | 121 (37%) |
|  | Complex | ≥ 1 day of additional training, or requires certification to achieve competency | 17 (7%) | 4 (4%) | 21 (6%) |

Seventy-two percent of observed RT-use (n=237) were for interventions. Most of these observed intervention RT had alternating current (AC) power (183, 78%), actuation (185,78%), no monitor (162, 68%), multi-use (222, 94%), and required a short training period (1-2 hours) for clinicians to establish competence (111, 47%). We observed PTs (173, 73%) using RT for interventions more than OTs (n=56, 24%). These devices saw the most use during one-on-one treatments (169, 71%). The most common type of intervention RT was balance and/or gait (n=93, 39%) followed by strength and/or endurance training (n=70, 30%; fig. 1).

**Figure 1:**
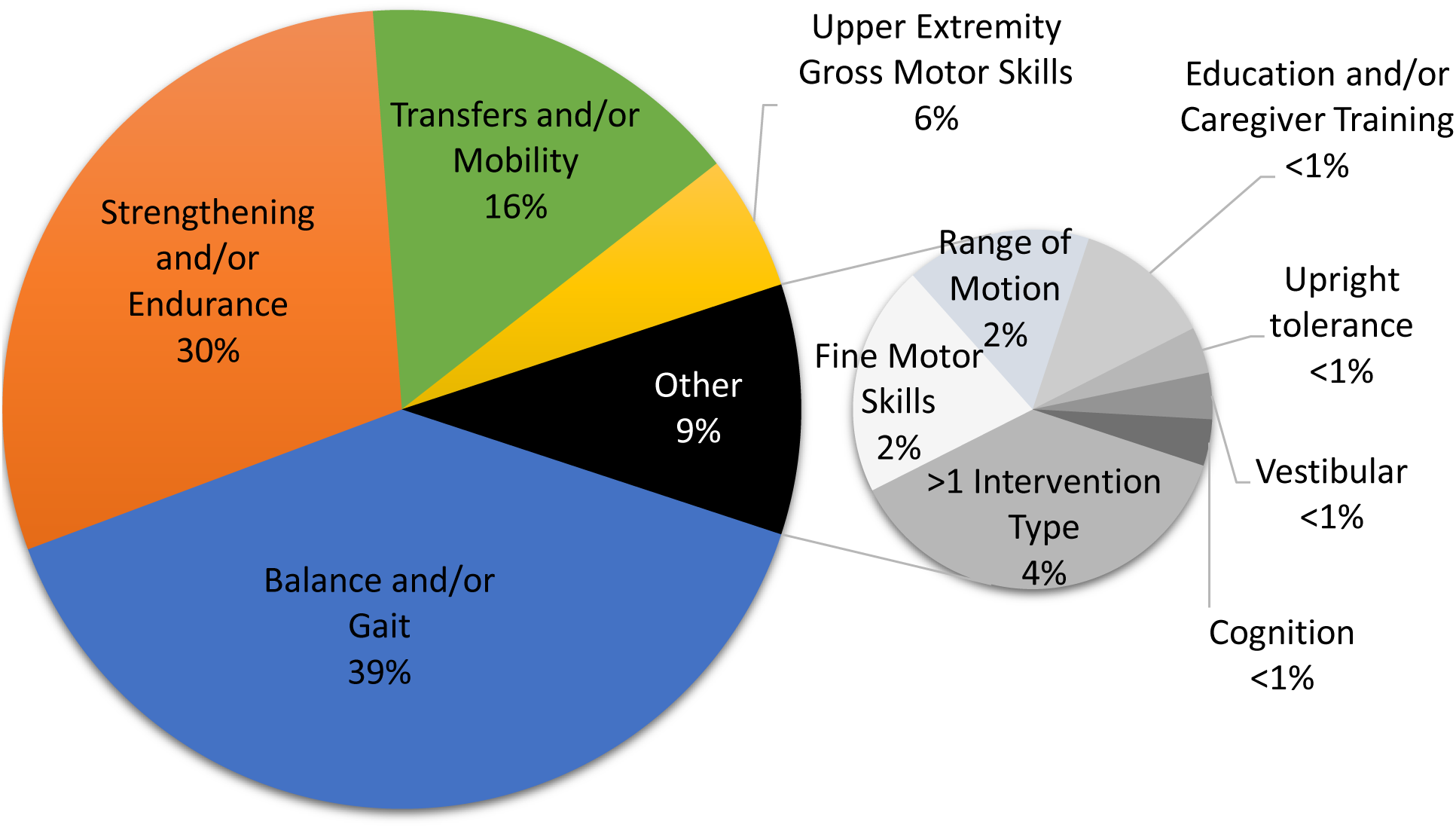
Intervention RT (n=237) subcategories. Devices for balance and gait (n=93, 39%) were the most common interventions observed, followed by devices for strengthening and/or endurance (n=70, 30%), and transfers and/or mobility (n=37, 16%).

Measurement RT accounted for 28% (n=92) of observed RT-use. Most of these observed measurement RT were battery-powered (81, 88%), unactuated (91,99%), had a monitor (81, 88%), had a single-use (90, 98%), and required only entry-level training (knowledge acquired in OT and PT school) (78,85%) -- this was because vitals (76, 83%) was the most common measurement type. We observed PTs (73,79%) using RT for measurement more than OTs (14,15%). The second most common measurement type was grip strength (6, 7%) followed by upper extremity function (5, 5%; fig. 2), both performed exclusively by OTs.

**Figure 2:**
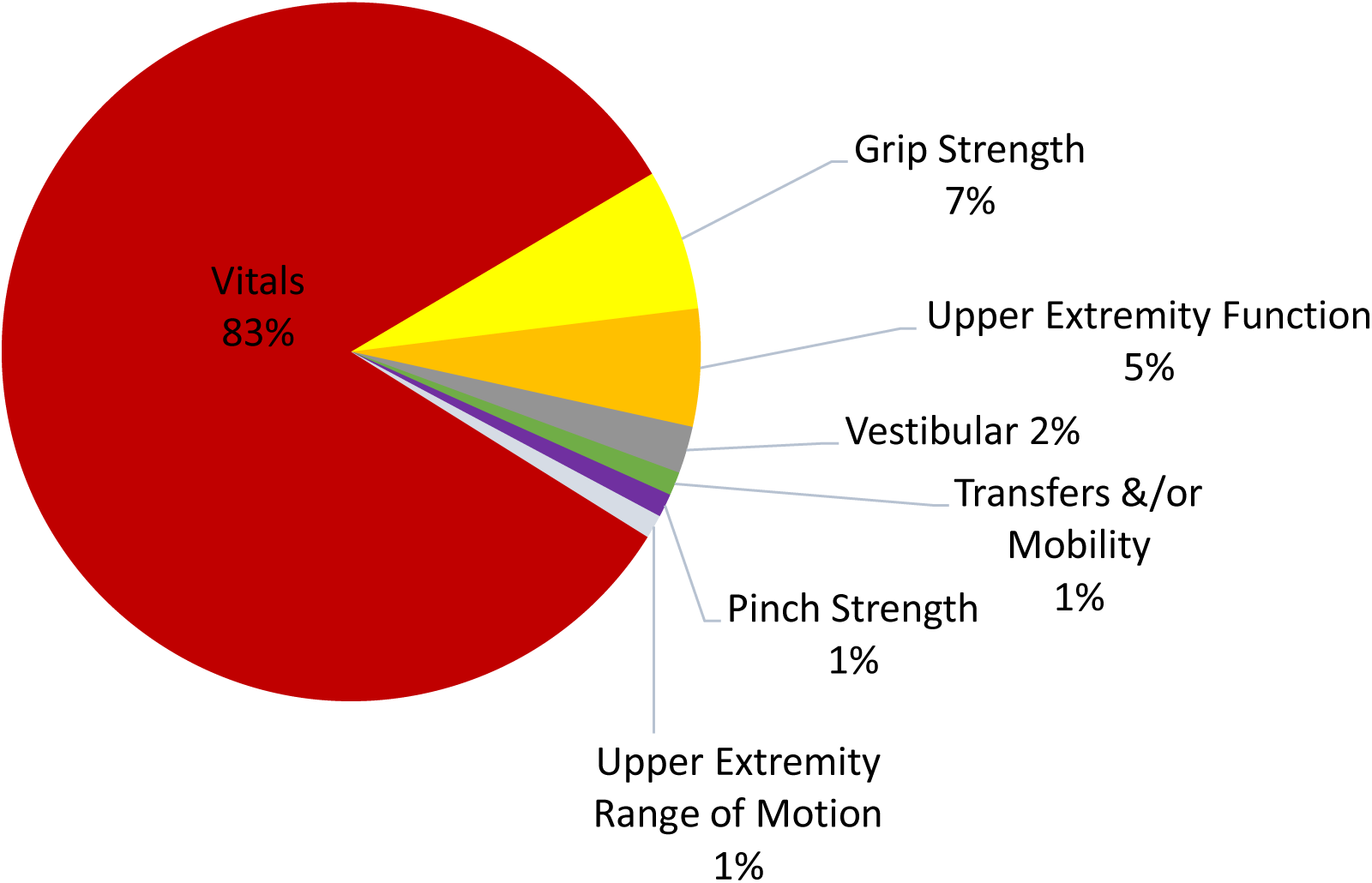
Measurement RT (n=92) subcategories. Vitals (n=76, 83%) dominated the observed measurements, followed by grip strength (n=6, 7%) and upper extremity function (n=5, 5%).

### Exploratory Data Analysis

We conducted exploratory data analysis to identify prevalent combinations of RT characteristics. We excluded non-novel RT, like the adjustable mat table (49, 15%) and adjustable parallel bars (24, 7%), to prevent bias.

For intervention RT, the dominant combination of characteristics included AC power, actuation, no display monitor, multi-use, requiring short training for competency, used during interventions targeting balance and/or gait, and used by PTs (40, 24%). These include body weight supported treadmills (n=38), overhead gait tracks with electric harness (n=2), and ceiling lifts (n=1). Excluding these, the second most common combination included unpowered, unactuated, no display monitor, multi-use, requiring short training for competency, used during interventions targeting balance and/or gait, and used by PTs (18,11%). The only devices in this category were manual over-head gait tracks (n=18).

For measurement RT, the dominant combination of characteristics included battery power, no actuation, with display monitor, tailored for single-use purpose, necessitating entry-level competency, and used by PTs (70, 97%). But vital signs monitoring machines accounted for all instances of the indicated measurement characteristics. Excluding vital signs, the dominant combination shifted to unpowered, unactuated, no monitor, tailored for single-use purpose, required short training for competency, and used by OTs (6,40%). These instances encompassed kits assessing upper extremity function, manual dexterity, and pinch/grip strength.

### Time

Figure 3 displays setup and total time spent with RT. Setup time was skewed, with a median of 1.2 minutes and a mean of 2.9±4.3 minutes. 84% of observations were of RT that took less than 5 minutes to setup, and 47% of RT took less than 1 minute to setup. In 5.4% of the observations, OTs and PTs used RT that took 10-36 minutes to set up. The RT that took the longest setup time were a mobile arm support that a patient purchased, and a clinician installed on the patient’s wheelchair (36 minutes), and an exoskeleton device (32 minutes). The average total time spent with RT (including setup and breakdown) was 20.5±16.3 minutes. Clinicians spent more time setting up and using intervention RT (setup mean 3.8±4.21 minutes; total time mean 25.6±15 minutes) compared to measurement RT (setup mean 0.8±1.3 minutes; total time mean 7.3±11.2 minutes).

**Figure 3:**
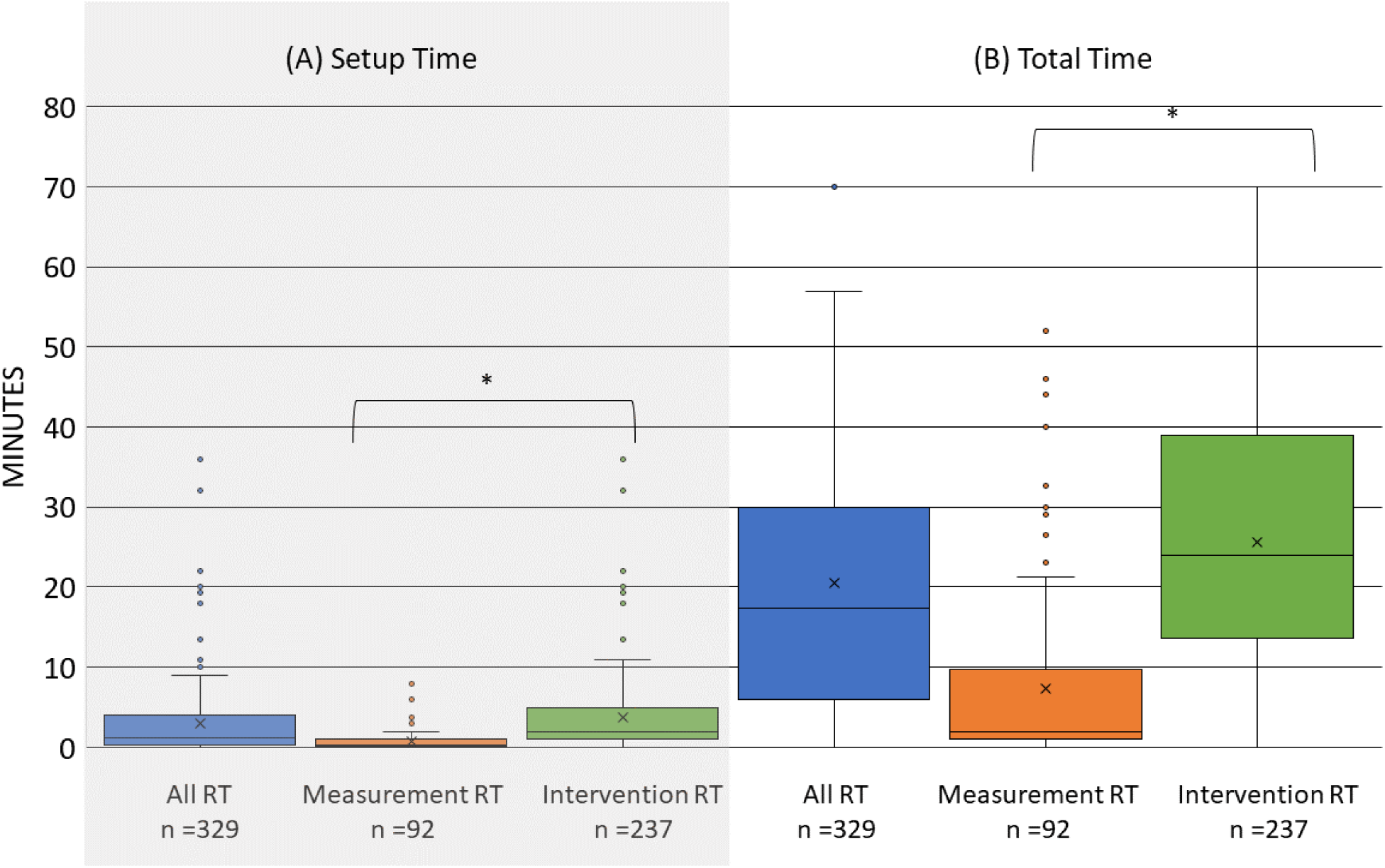
Setup time (panel A) and total time (panel B) spent with RT devices among OTs and PTs during observed sessions. More time is spent setting up and using RT for intervention than measurement (*p<0.001).

### Therapists’ Behaviors

We investigated clinicians’ instructions and patients’ experience levels with RT. Predominately, therapists used verbal instructions (313, 72%) when using RT, followed by a combination of verbal and demonstrated instruction (46, 14%). Most patients were repeated RT users (294, 89%) rather than first-time users (35, 10.6%). First-time users received verbal instructions or a mix of verbal and physical instruction (e.g., demonstration, physical cues, physical assistance). Among the patients with no instructions (16, 4.9%) all were repeat users, using vitals machines or exercise equipment.

We documented clinicians’ feedback types as either knowledge of performance (task quality or movement patterns), or knowledge of results (outcomes), based on a commonly-used distinction in motor learning research.^25–27^ Instances of no feedback were rare (4, 1.2%). Clinicians primarily provided knowledge of performance (197, 59.7%) compared to knowledge of results (99, 30%) or a combination of both (30, 9%).

Our findings revealed that clinicians were evenly divided between providing one-on-one direct attention (164, 49.7%) and multi-tasking (166, 50.3%) when treating patients with RT.

## DISCUSSION

We aimed to understand RT usage patterns such as the characteristics of RT, time spent on setup and total use, and therapists’ behaviors.

### Rehabilitation Technology Characteristics

Clinicians embraced smart, actuated (185, 78%), and AC-powered (183, 77%) intervention RT, often observed during balance and/or gait training (93, 39%), including body weight supported treadmills (n=38), used by physical therapists (173, 73%). The preference may stem from the safety and efficacy benefits of body weight support technology in gait training.^15^

The inventory revealed a diverse range of RT for balance and/or gait training, but fewer than half were observed in used. Despite evidence to support split-belt treadmill training,^28,29^ and emerging research advocating for exoskeletons,^30^ both devices were rarely observed in use. The “relative advantage” barrier, identified in our previous work, persists as there is no clear advantage of novel RT over existing alternatives,^19,31^ and clinicians favor familiar devices such as body-weight supported treadmills and over-head gait tracks due to their proven effectiveness. This preference is mirrored in the hospital system, with each inpatient floor and setting equipped with at least one body weight supported treadmill and gait track; split-belt treadmills and exoskeletons are limited to 1-2 in the entire system. This highlights the need for developers of RT to align their devices with clinician practices, workflow, and practice guidelines. Novel RT needs to demonstrate a clear benefit over the existing equipment.

While intervention RT outnumbered measurement RT, more measurement devices in the inventory were observed in use with the most used measurement RT being vital machines and grip strength dynamometers. Vitals measurement is essential for PTs to monitor patient safety in all conditions, and aerobic intervention efficacy; plus, they are faster and more accurate than taking manual measurements.^32^ Grip strength dynamometers are easy to use, provide more precise measurement than the alternative manual muscle testing, and have established psychometric properties in nearly all conditions.^33,34^ Two important features of these successful measurement RT are their adaptability to different populations and their relative advantage over prior gold standard measures of the vitals and strength domains.^31^ Developers are likely to succeed by enhancing precision in current clinical metrics, instead of expending resources on the creation of novel measurement criteria.

### Time Constraints

Observed RT typically requires less than a day’s worth of additional training, and clinicians did not invest a lot of time in setting up RT. This aligns with our earlier work: the time it takes to learn and set up RT is an important factor in its uptake.^19^ Clinicians exhibit a stronger preference for RT that is integrated into their entry-level academic training,^19^ and constraints on learning during work hours, coupled with the desire to optimize patient treatment time, discourage extensive setup.^13^ We advise developers to consider training and setup time in RT development, even for devices with high efficacy and effectiveness. While greater efficacy might encourage more time investment, our field observations offer a benchmark for designing user-friendly RT.

### Therapists’ Behaviors

Feedback plays a crucial role in motor learning, and clinicians’ choice of feedback can influence how much patients learn.^25^ Our observations revealed a preference among clinicians for providing knowledge of performance rather than knowledge of results when using RT. These findings align with current stroke research suggesting that knowledge of performance results in superior motor learning compared to knowledge of results^26,27^ and highlight therapists’ attention to movement quality. Future research should investigate if clinician feedback type differs in the presence or absence of RT, or examine the correlation of specific types of feedback (directed vs guided cuing) on patient outcomes. Ultimately, personalized feedback types and frequencies may be necessary based on the patient’s condition.^27^

Adhering to documentation standards dictated by insurance and legal requirements ^35,36^ often imposes an administrative burden contributing to clinician burnout.^37^ Allowing patients to independently work with a device under clinical oversight provides therapists with valuable hands-free moments for efficient documentation. Our observations indicate no clear preference towards multi-tasking during RT use, suggesting potential RT solutions that facilitate engaging with patients or multi-tasking between other administrative demands and patient care.

### Study Limitations

Interpretations of the results can serve different purposes: they can reveal characteristics of RT associated with common use, which might guide RT designers, or highlight potential biases associated with lack of use, which might inform implementation innovators. Restricting our data collection to only the treatment gyms is a limitation, since there are devices located in managers’ offices, patient rooms, and on research floors. This leads to underreporting of RT-use during clinical treatment and limits the scope of the RT inventory. The 90 available technologies in the inventory are likely larger than many smaller rehabilitation settings but were still insufficient in size for advanced statistical modeling. We based the categorization of RT on observed use rather than the inventory, which has the potential of introducing bias. It is important to note that we completed our RT categorization at one organization, and it is not exhaustive; thus, it may not encompass the full spectrum of clinical treatment. Finally, our reliance on observation with little interactions with the clinicians makes it difficult to extrapolate the underlying reasons for specific clinical behaviors. These limitations highlight the need for further research on motivations behind RT choices within the health system.

## CONCLUSIONS

This mixed methods field observation study in a technology-focused hospital revealed that majority of available RT was not observed used. We observed intervention RT used twice as frequently as measurement RT, with a focus on gait/balance training and strengthening. The measurement RT that is used is primarily for vitals followed by grip strength. These results highlight the ongoing struggle to integrate new RT in clinical practice: successful cases are rare and involve well-established technologies like treadmills, exercise bicycles, and grip strength sensors. The limited use of technology for measurement is notable. Also, used RT tend to have brief set-up times (less than 4 minutes), and, when using them, clinicians typically provide a lot of feedback, focusing on performance (e.g., movement quality) rather than results. In about half of use cases, clinicians leverage the time the patient is interacting with technology to work on documentation.

## Supporting information

Supplemental Interactive Sankey Diagram

## Data Availability

All data produced in the present study are available upon reasonable request to the authors

https://sralab-kteam.shinyapps.io/rt_analysis/

## ACKNOWLEDGEMENTS

We thank the clinical staff at the Shirley Ryan AbilityLab (SRALab) for allowing us to perform the field observations. We also thank the Robotics Lab and KTEAM at SRALab and the members of the STARS RERC and COMET-RERC.org for their ongoing comments and advice.

This study was presented as a poster at Progress in Clinical Motor Control II Movement and Rehabilitation Sciences conference at Northwestern University in Chicago, IL , on July 13^th^, 2023.

## ABBREVIATIONS

(OT): Occupational Therapy
(PT): Physical Therapy
(RT): Rehabilitation Technology

## SUPPLIERS

a. Excel spreadsheet by Microsoft.
b. R Statistical Software; R Foundation for Statistical Computing.
c. Python Software Foundation (Libraries Used: Pandas).

